# A qualitative exploration of the perspectives of international medical students residing in university hostels amid COVID-19 pandemic lockdown

**DOI:** 10.1101/2021.08.02.21261480

**Authors:** Sitaram Khadka, Muhammad Usman, Mohammad Saleem, Moshin Ali, Huma Rasheed, Santoshi Giri, Hafiz Asad Saeed, Ravi Prasad Gupta, Yogesh Bajgain, Janak Shahi

**Author notes:** Name: Muhammad Usman, Contact, Address: Institute of Pharmaceutical Sciences, University of Veterinary and Animal Sciences, Lahore, Pakistan.

## Abstract

**Background:** COVID-19 pandemic has portentously frightened the existence of life all over the world. The lockdown approach was adopted as a containment strategy as the disease itself has manifested severe social, economic, and psychiatric implications.

**Objectives:** To explore the perception and preparedness of international medical students residing in university hostels amid the COVID-19 pandemic lockdown.

**Design:** A semi-structured interview guide was developed in this qualitative study design. All the interviews were audio-taped, transcribed verbatim, and then analyzed for thematic contents by standard content analysis framework.

**Setting:** Interviews were conducted in university hostels in Punjab, Pakistan.

**Participants:** A total of 11 international medical students were interviewed face-to-face through the purposive sampling technique to obtain in-depth individual viewpoints.

**Results:** The thematic content analysis yielded five major themes: Familiarity with COVID-19, Perceptions and attitudes towards COVID-19, Preparedness for safety against COVID-19, Barriers to lifestyle, and Psychological perspectives. A better general perception and preparedness among international medical students regarding COVID-19 was found. Good knowledge regarding the overview of COVID-19; adequate preventive approaches such as social distancing, use of masks, gloves, and sanitizers; and compliance with the lockdown measures were reported by the respondents. The pertinent issue raised by the respondents is the disturbance in normal routine due to distortion in social life and isolation that may cause psychological stress.

**Conclusions:** The findings from this study lighten the people’s perspectives that help the government to prepare public health strategies based on population-focused approaches. The present study demonstrates the respondents’ opinion on COVID-19 management by personal hygiene, social distancing, and complying with the lockdown measures. Furthermore, it demands that timely and evidence-based teaching-learning techniques should be adopted for students’ engagement which ensures mental health and self-motivation as well. Therefore, they can utilize their time productively which could have a long-term effect on their careers and healthcare services.

## INTRODUCTION

On 11 March 2020, the World Health Organization (WHO) declared COVID-19 as a global pandemic ever since it has manifested itself as serious public health and developmental challenge that has viciously affected all the domains of quality of life including health, socio-economic, and education.^1^ Till date, it has affected more than one hundred and ninety-five million people with more than four million deaths around the world.^2^ Even after the passage of a year, the uncertainty of the approved therapeutic agent exists for the therapy of COVID-19. However vaccines are now being available recently for people aged older than 12 years, it still takes more than a year to fully cover all the countries around the globe.^3,4^ In such adverse scenarios, repurposing of prior available medications and supportive treatment is being practiced as a treatment option considering all the pros and cons.^5^ Various preventive approaches such as lockdown, social distancing, adequate hygiene and sanitation, and wearing masks for breaking the chain of transmission are the mainstay of patient care as of now.^6^

Though the international students are not enlisted in a high-risk group, they are severely affected during the lockdown period socially and economically given their immigration status.^7^ International students, particularly from lower and middle-income countries (LMICs) are already facing challenging circumstances including financial crunches. Lockdown measures add to their plight and potentially lead them into social and psychological stress.^7^

Pakistan takes students from different parts of the world for higher education in the medical and non-medical fields. Most of the international students stay in hostels provided by their universities. Amid the COVID-19 pandemic, they are confined to the hostels. Disruption in education, travel restriction, closure of the mess, isolation, closed border amid lockdown have impaired their daily activities. Fear of infection, anxiety, hoax calls, cyberbullying, and financial crisis may cause a substantial impact on mental health amid COVID-19 preventive activities.^8-10^ Such stress-associated disorders may lead to suicidal behavior as well.^11^ In this scenario, assessing the stress level of international students who are confined in hostels amid the COVID-19 pandemic and understanding their perception and preparedness aspect regarding the COVID-19 pandemic is crucial. Such information can be a milestone to make concerned authorities aware of adverse consequences that may arise in case of obliviousness in managing their daily activities.

This study has been conducted to answer the following question: What are the perceptions and preparedness of international medical students residing in university hostels amid the COVID-19 pandemic lockdown in Pakistan?

To the best of our knowledge, this is the first of its kind study designed to evaluate the challenges faced by international medical students residing in university hostels amid COVID-19 lockdown and their perceptions and preparedness towards it which might help the concerned authorities to prepare relevant public health strategies.

## METHODS

### Ethical approval

The ethics approval was received from the Institutional Ethics Review Board of the University of the Punjab, Lahore, Pakistan (Reference number:183/DFEMS). Informed verbal consent to participate in the study was obtained from participants.

### Study design

A qualitative research methodology was adopted as it is flexible and focuses on non-numeric data that consent to an in-depth understanding of the respondent’s perspective. Moreover, qualitative analysis supports identifying and fill in gaps that are left unnoticed by the quantitative types.

The COREQ (consolidated criteria for reporting qualitative research) checklist was utilized for reporting qualitative studies.^12^

### Instrument development

A semi-structured interview guide was developed. It was tested for validity and reliability by two skilled researchers at the University of Veterinary and Animal Sciences, Lahore, Pakistan (B and E). It was pre-tested and verified for accuracy and consistency. The reliability of the findings was assured by recording face-to-face interviews with the international students following their approval.

### Respondents sampling and inclusion criteria

The study was conducted in Punjab Province, which is the most developed, populated, and the second-largest province of Pakistan. The participants in the study were recruited using the purposive sampling technique based on their availability until saturation was achieved. A total of 11 international medical students residing in university hostels were finally selected for interview. The targeted participants were international students from various faculties of medicines such as pharmacy, veterinary, medical, and dental in universities around Punjab Province, Pakistan.

### In⍰depth face⍰to⍰face interviews

After giving an explanatory statement with the study objectives to the participants, verbal consent was taken. To ensure privacy, the identity of the respondents was kept confidential and the anonymity of opinions was also confirmed by using codes by the two researchers (SK and MU). Interviews were continued until the saturation point from where new data doesn’t occur. The respondents were aware of voluntary involvement in the interview, and the independence to drop the participation at any time. The co-authors of the study interviewed the participants in their hostel room between April 2020 and June 2020 in the English language. All the interviews were audio-taped, and additional field notes were taken by the principal investigator SK (male). Suitable probing was done to seek out more information. The respondents were free to describe additional views on the topic. The duration of each interview was approximately 20-30 minutes.

### Analysis

All the recorded interviews were transcribed verbatim. The comments were changed to some extent for grammatical corrections during the data extraction process. Based on the method described by Braun and Clarke, the data were thematically analyzed manually by in-depth penetration into the interviews.^13^ The significant answers were deductively highlighted. Then the answers were compared and contrasted to establish any variances and themes were identified by successively reorganizing the pattern. The transcribed interviews were analyzed for interconnected comments while categorizing the theme. This exercise helped cluster the statements that were previously considered unrelated or discrete. The interview was conducted in a very inductive and flexible manner to further explore the valid issues.

## RESULTS

### Demographics of the participants

A total of 11 participants from seven different Universities in Pakistan were included in the study (n=11). The majority of them were from Universities located in Lahore (n=8) followed by Multan (n=2), and Faisalabad (n=1). The participants belonged to six different countries of the Eastern Mediterranean Region (n=3), South-East Asia Region (n=2), and African Region (n=1). There was a total of eight male students and the rest were female among the participants. The majority of the participants (n=8) were of more than 25 years of age. Under-graduate students were nine in number followed by post-graduate level students. All the participants were staying in Pakistan for more than one year. The majority of the participants (n=6) were from pharmacy faculty, three from medical, and one each from dentistry and veterinary faculties.

A flow diagram of the participants’ recruitment is given in **Figure 1**.

**Figure 1.**
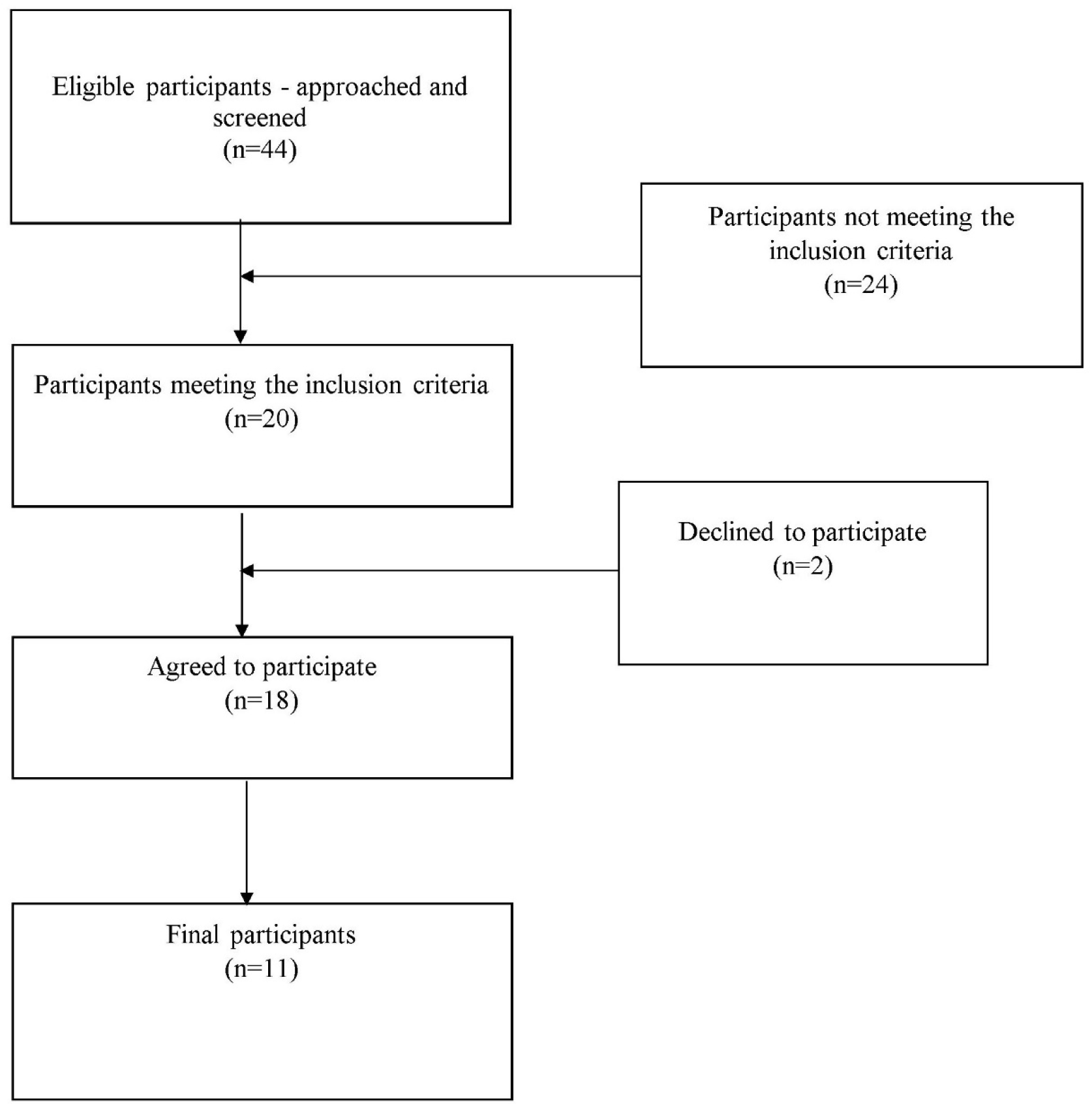
A flow diagram of the participants’ recruitment for the qualitative interviews

The demographic distribution of the participants is described below in **Table 1**.

**Table 1.** Demographics of the participants

| Characteristics | Parameters | Frequency (n) | Percentage (%) |
| --- | --- | --- | --- |
| Gender | Male | 8 | 72.72 |
|  | Female | 3 | 27.27 |
| Age | <25 years | 8 | 72.72 |
|  | >25 years | 3 | 27.27 |
| Education (Level) | Under-graduate | 9 | 81.82 |
|  | Post-graduate | 2 | 18.18 |
| Education<br>(Discipline) | Medicine | 3 | 27.27 |
|  | Pharmacy | 6 | 54.55 |
|  | Dentistry | 1 | 9.09 |
|  | Veterinary science | 1 | 9.09 |
| Stay duration | <1 year | 0 | 0 |
|  | >1 year | 11 | 100 |
| Country of origin | Nepal | 6 | 54.55 |
|  | Sri Lanka | 1 | 9.09 |
|  | Syria | 1 | 9.09 |
|  | Yemen | 1 | 9.09 |
|  | Iran | 1 | 9.09 |
|  | Kenya | 1 | 9.09 |

### Thematic analysis of the content

The thematic analysis of the content of the interview led to five major themes: 1. Familiarity with COVID-19 (Knowledge), 2. Perceptions and attitudes towards COVID-19 (Attitude), 3. Preparedness for safety against COVID-19 (Practice/Action), 4. Barriers to lifestyle (Experience/Outcome), and 5. Psychological perspectives (Experience/Outcome).

### Theme 1: Familiarity with COVID-19

#### Knowledge about COVID-19 pandemic - Overview

The participants were interviewed regarding their knowledge about the COVID-19 pandemic. All of them answered in a similar way regarding the nature of the disease, origin, and causative agent, Table 2.

**Table 2.**
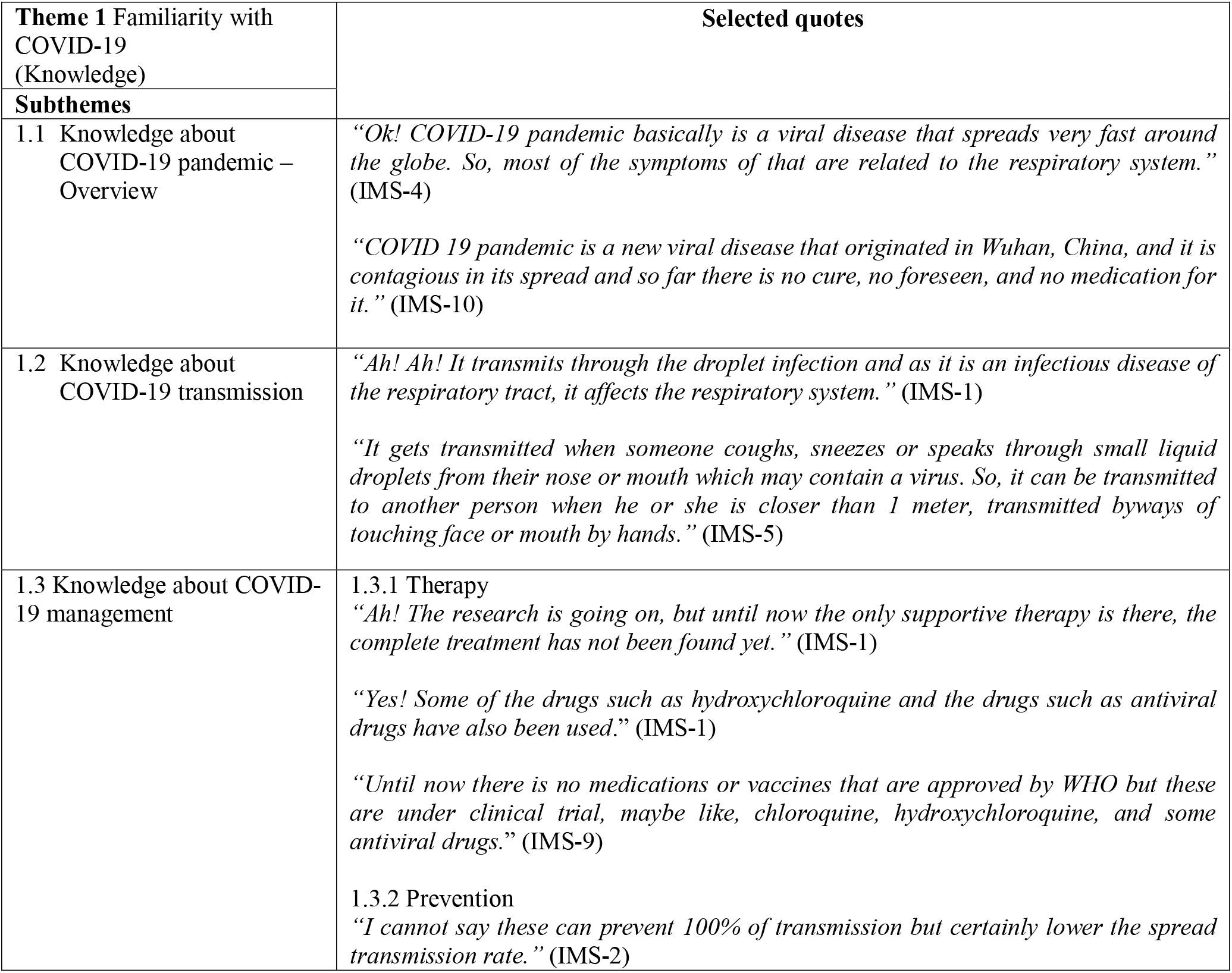
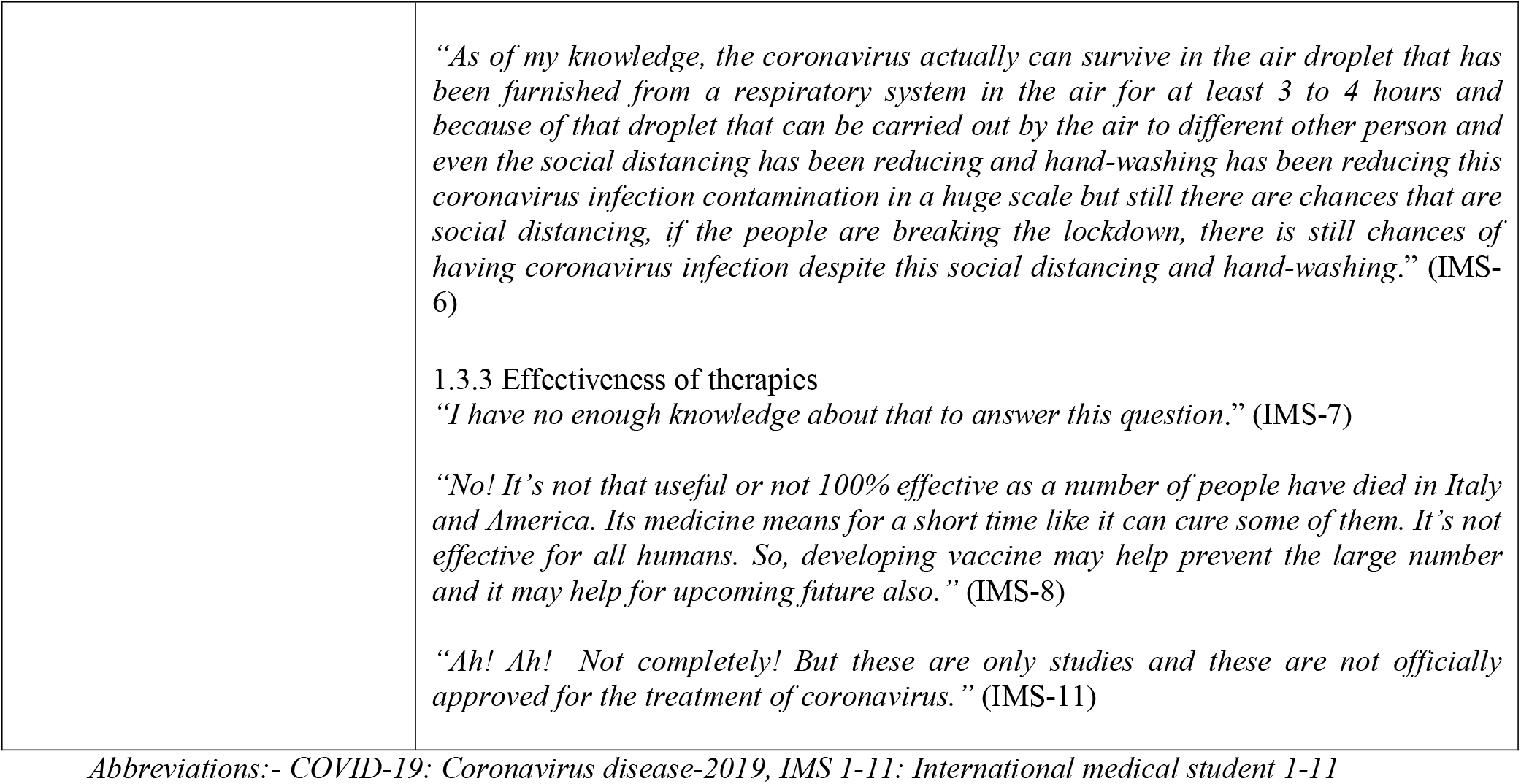
Thematic analysis of the interview data and selected quotes of the respondents (Theme 1)

| Theme 1 Familiarity with COVID-19 (Knowledge) | Selected quotes |
| --- | --- |
| Subthemes |  |
| 1.1 Knowledge about COVID-19 pandemic – Overview | *“Ok! COVID-19 pandemic basically is a viral disease that spreads very fast around the globe. So, most of the symptoms of that are related to the respiratory system.” (IMS-4)*<br>*“COVID 19 pandemic is a new viral disease that originated in Wuhan, China, and it is contagious in its spread and so far there is no cure, no foreseen, and no medication for it.” (IMS-10)* |
| 1.2 Knowledge about COVID-19 transmission | *“Ah! Ah! It transmits through the droplet infection and as it is an infectious disease of the respiratory tract, it affects the respiratory system.” (IMS-1)*<br>*“It gets transmitted when someone coughs, sneezes or speaks through small liquid droplets from their nose or mouth which may contain a virus. So, it can be transmitted to another person when he or she is closer than 1 meter, transmitted byways of touching face or mouth by hands.” (IMS-5)* |
| 1.3 Knowledge about COVID-19 management | **1.3.1 Therapy**<br>*“Ah! The research is going on, but until now the only supportive therapy is there, the complete treatment has not been found yet.” (IMS-1)*<br>*“Yes! Some of the drugs such as hydroxychloroquine and the drugs such as antiviral drugs have also been used.” (IMS-1)*<br>*“Until now there is no medications or vaccines that are approved by WHO but these are under clinical trial, maybe like, chloroquine, hydroxychloroquine, and some antiviral drugs.” (IMS-9)*<br>**1.3.2 Prevention**<br>*“I cannot say these can prevent 100% of transmission but certainly lower the spread transmission rate.” (IMS-2)* |
|  | *“As of my knowledge, the coronavirus actually can survive in the air droplet that has been furnished from a respiratory system in the air for at least 3 to 4 hours and because of that droplet that can be carried out by the air to different other person and even the social distancing has been reducing and hand-washing has been reducing this coronavirus infection contamination in a huge scale but still there are chances that are social distancing, if the people are breaking the lockdown, there is still chances of having coronavirus infection despite this social distancing and hand-washing.” (IMS-6)*<br>1.3.3 Effectiveness of therapies<br>*“I have no enough knowledge about that to answer this question.” (IMS-7)*<br>*“No! It’s not that useful or not 100% effective as a number of people have died in Italy and America. Its medicine means for a short time like it can cure some of them. It’s not effective for all humans. So, developing vaccine may help prevent the large number and it may help for upcoming future also.” (IMS-8)*<br>*“Ah! Ah! Not completely! But these are only studies and these are not officially approved for the treatment of coronavirus.” (IMS-11)* |

Abbreviations:- COVID-19: Coronavirus disease-2019, IMS 1-11: International medical student 1-11

#### Knowledge about COVID-19 transmission

The responses of all participants were almost similar regarding the modes of transmission, Table 2.

#### Knowledge about COVID-19 management

The majority of the participants had an appropriate concept about the therapy of COVID-19. Some of the participants were a little bit confused regarding the name of the drugs used as a treatment option. However, many of them had an appropriate concept about ongoing trials for therapeutic and prophylactic agents and the use of only supportive therapy and the best possible option of treating agents available for COVID-19 management.

##### Therapy

The respondents’ views are provided in table 2.

##### Prevention

Answering a query regarding social distancing, hand washing, using masks, gloves, and goggles as preventive measures; a majority of the respondents stated that these can only lower the transmission rate but cannot prevent it at 100%. Some of the participants believed that these measures help in the complete prevention of transmission of COVID-19. Different students expressed various views about the availability and survival of coronavirus in different mediums, Table 2.

##### Effectiveness of therapies

Though the majority of the students reported that the adopted treatment methods are only being used as the best possible option, some of them were not being able to express a clear answer regarding the effectiveness of the ongoing therapy, Table 2.

### Theme 2: Perception and attitudes towards COVID-19

#### Risk of contracting COVID-19

Regarding the risk of contracting COVID-19, all of the participants considered that they are at risk but some participants claimed that they are in a safer state than any other persons as they were confined in the hostels, Table 3.

**Table 3.** Thematic analysis of the interview data and selected quotes of the respondents (Theme 2)

| **Theme 2** Perceptions and attitudes towards COVID-19 (Attitude) | **Selected quotes** |
| --- | --- |
| **Subthemes** |  |
| 2.1 Risk of contracting COVID-19 | *“Especially we are in a hostel, if the sweeper or colleague or anyone inside the hostel, they are affected, you know, by corona, is easily transmitted from them.” (IMS-3)*<br>*“Yes! I think there is a possibility. Yes, there will be a possibility but any of the students in the hostel get infected. So, I think, then we all will get infected but we are safer than others outside the campus.” (IMS-4)* |
| 2.2 Community level-Lock down approach | *“Lockdown is very important and effective for preventing a spread of COVID-19.” (IMS-5)*<br>*“Yes! It is an effective way to stay at home and it has, kind of, prevented spread like it has broken spread of coronavirus, like, it’s less infective. There was more possibility that people may get suffer but due to lockdown, fewer people are suffering and we are kind of safe.” (IMS-8)* |
| 2.3 Institutional level- University/hostel management | *“I think this is not good at facilities.” (IMS-11)*<br>*“Yes! Good! Right! Right!” (IMS-9)*<br>*“Ah! There are also precautionary measures and all the sanitary measures are satisfactory but not 100% perfect but our continuous effort is to warn the administration for the better facilities provided here.” (IMS-2)* |
| 2.4 Personal level- Tackling aspects | *“Umm! If my hostel fellow gets affected by COVID-19, then he should go for isolation. We must be staying in quarantine and, ah! Those we who are in contact with him should stay in isolation too or might be quarantined.” (IMS-1)*<br>*“Umm! First of all, I will call my hospital emergency and say about my conditions, and say these are happening, and according to that situation they will tell me what to do further.” (IMS-1)*<br>*“I think, there are rules set even when one shows such kind of symptoms, he should isolate himself, and of course, I will also have to take the precautionary measures like not coming in touch with the infected.” (IMS-10)*<br>*“Ok! What I will do is, I will report. You know there is a number I’m supposed to call if I show such kind of symptoms. I will report and then, from there, I will take the instruction they will give me” (IMS-10)* |
| 2.5 Perception to rules and guidelines | *“Yes! I believe every individual is responsible for this condition, responsible for following the necessary guidelines for the prevention of this disease. Because this is not only yourself who is in danger, there is our family and the whole people of the world are in danger. So, we must do whatever we are responsible for to protect ourselves and the whole world.” (IMS-11)*<br>*“Umm! Yes! I want to comment that let’s hope for the best and let’s follow the guidelines that are provided by the local authorities and our government, and let’s stay strong. We will get through this.” (IMS-1)* |

Abbreviations:- COVID-19: Coronavirus disease-2019, IMS 1-11: International medical student 1-11

#### Community level-Lock down approach

Regarding the lockdown approach as a containment strategy to prevent COVID-19, all the respondents showed a positive response, Table 3.

#### Institutional level-University/hostel management

The mixed views were obtained upon asking for university and hostel management amid the COVID-19 pandemic. Some of the respondents even indicated dissatisfaction about the local scene as the people were not taking the lockdown seriously, Table 3.

#### Personal level -Tackling aspect

Upon a question related to tackling COVID-19 in case their fellow hostelers get infected with COVID-19, most of them asserted that they would suggest isolation and quarantine, while some of the respondents were intending to take them to hospital, while others did not state about isolation, quarantine, but discussed supportive therapy and other laboratory tests to conduct.

While replying to the query “What will you do if symptoms of flu that resemble COVID-19 occur?”, the respondents emphasized on calling an emergency number, some of them were intending to get the test done and isolate themselves, while others said that they follow preventive measures to tackle such scenario, Table 3.

#### Perception to rules and guidelines

All of the respondents were found positive towards complying with the rules and guidelines set for the management of such a global health threat in an optimistic way, Table 3.

### Theme 3: Preparedness for safety against COVID-19

#### Preventive measures

Regarding preventive measures adopted against the COVID-19 pandemic, it was found that they carry hand sanitizer, wear masks and gloves, usually wash their hands with soap water, and follow social distancing as per guidelines, Table 4.

**Table 4.**
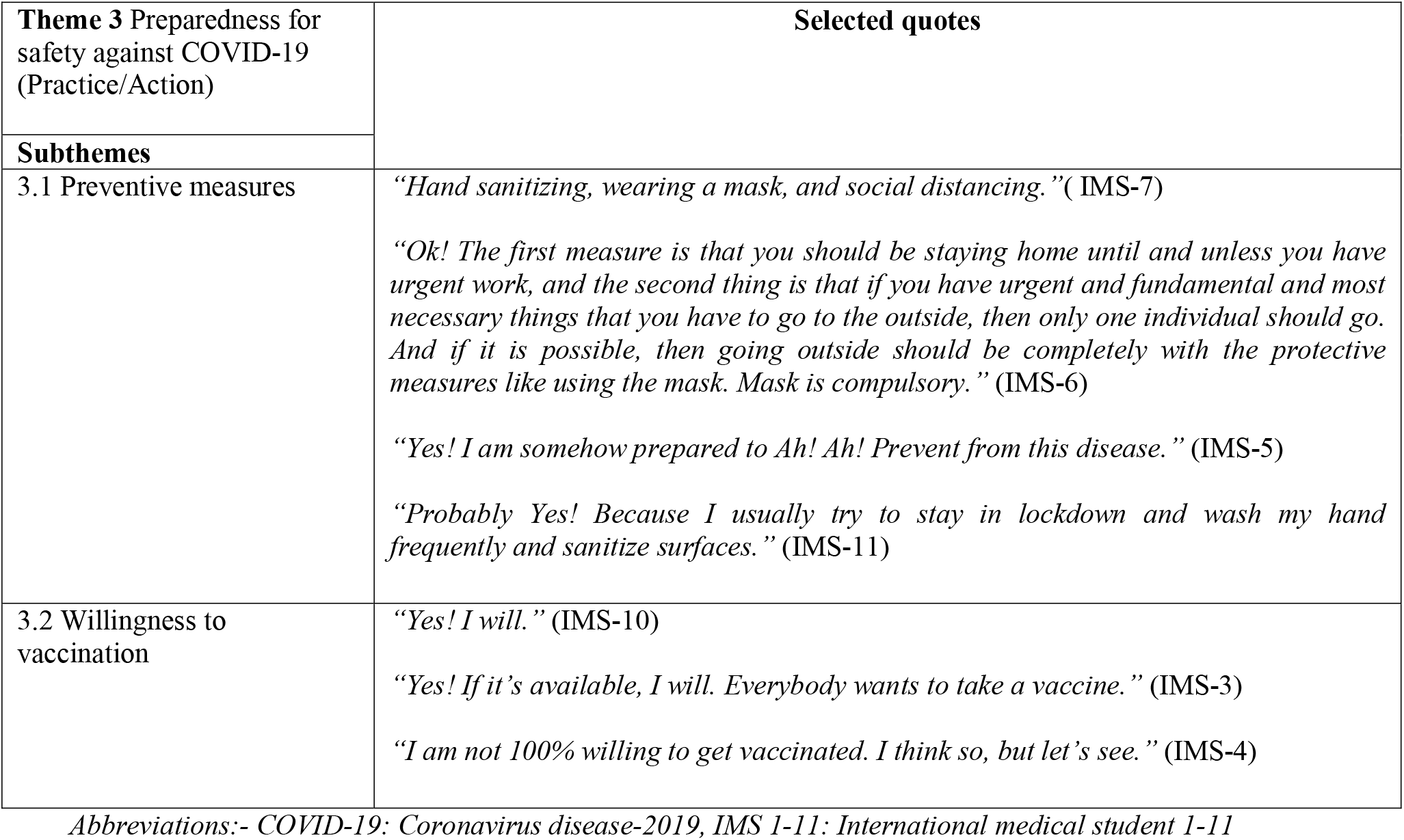
Thematic analysis of the interview data and selected quotes of the respondents (Theme 3)

| **Theme 3** Preparedness for safety against COVID-19 (Practice/Action) | **Selected quotes** |
| --- | --- |
| **Subthemes** |  |
| 3.1 Preventive measures | *“Hand sanitizing, wearing a mask, and social distancing.”( IMS-7)*<br>*“Ok! The first measure is that you should be staying home until and unless you have urgent work, and the second thing is that if you have urgent and fundamental and most necessary things that you have to go to the outside, then only one individual should go. And if it is possible, then going outside should be completely with the protective measures like using the mask. Mask is compulsory.” (IMS-6)*<br>*“Yes! I am somehow prepared to Ah! Ah! Prevent from this disease.” (IMS-5)*<br>*“Probably Yes! Because I usually try to stay in lockdown and wash my hand frequently and sanitize surfaces.” (IMS-11)* |
| 3.2 Willingness to vaccination | *“Yes! I will.” (IMS-10)*<br>*“Yes! If it’s available, I will. Everybody wants to take a vaccine.” (IMS-3)*<br>*“I am not 100% willing to get vaccinated. I think so, but let’s see.” (IMS-4)* |

*Abbreviations:- COVID-19: Coronavirus disease-2019, IMS 1-11: International medical student 1-11*

#### Willingness to vaccination

Regarding vaccination, mixed views were obtained, Table 4.

### Theme 4: Barriers to the lifestyle

The lockdown approach and social distancing adopted as a containment strategy were considered barriers to the everyday lifestyle during the COVID-19 pandemic. They were not shaking hands, hugging friends, sharing stuff such as phones and utensils, and touching any surfaces outside the room which they felt unusual, though following these as a guideline, Table 5.

**Table 5.** Thematic analysis of the interview data and selected quotes of the respondents (Theme 4)

| Theme 4 Barriers to lifestyle | Selected quotes |
| --- | --- |

| (Experience/Outcome) |  |
| --- | --- |
| Subthemes |  |
| 4.1 Impact of movement restriction | *“Yes! Mainly my education system is changed. I am studying online.” (IMS-7)*<br>*“Yes! I was going back home but couldn’t because of a virus.” (IMS-9)*<br>*“I only go to buy necessary things, like, once in a week or twice maybe, sometimes to buy goods; the necessary goods, the food that is necessary.” (IMS-8)* |
| 4.2 Effect on daily activities | *“Ah! We used to go to the hospital. We used to do our job like 5 to 6 hours a day, but now we are going only 1 day per week, and ah! I also cannot go outside and we used to go outside for exercise. Ah! And it has also affected differently - our food, our way of eating, way of sleeping, everything is affected.” (IMS-1)*<br>*“Yes! Because it reduces the daily physical activities and forces us to do our work almost at home.” (IMS-11)*<br>4.2.1 Lifestyle dropped due to COVID-19<br>*“Ah! Yes! It changes lifestyle because we have to stay at a hostel and we can’t do daily activities like as usual.” (IMS -5)*<br>*“Yes! Definitely! For example, I don’t get to hug my friends. I don’t get to shake hands. I don’t get to go out as I used to do before when my studies, my studies are disrupted. So, completely it has changed my life.” (IMS-10)*<br>4.2.2 Lifestyle adopted due to COVID-19<br>*“I am just watching umm! YouTube videos, doing stuff and sometimes read books, and sometimes I see my phone like that.” (IMS-1)*<br>*“Currently, I am mostly using social media. Then in free time like you can say, after getting bored from social media, I read a book.” (IMS-2)*<br>*“Yes! I usually read news articles about COVID-19 and follow WHO.” (IMS-11)* |

Abbreviations:- COVID-19: Coronavirus disease-2019, IMS 1-11: International medical student 1-11

#### Impact of movement restriction

Not getting an opportunity for conventional classes, not going out from the hostel frequently, not being able to go home, and being confined within the hostel premises were among the responses obtained, Table 5.

#### Effect on daily activities

The majority of the respondents replied that changes in routine which disturbed timing to go for internship, physical activities, and meal timing were the most affected daily activities by the COVID-19 pandemic, Table 5.

##### Lifestyle dropped due to COVID-19

Regarding lifestyle changes due to the COVID-19 pandemic, the responses are depicted in Table 5.

##### Lifestyle adopted due to COVID-19

Pastime activities of most of the respondents during the COVID-19 pandemic were reading books including course books and news articles, playing games, and using social media, Table 5.

### Theme 5: Psychological perspectives

#### Fear of COVID-19

There were mixed reactions regarding fear of COVID-19 such as some were scared, while others were cautious rather than being scared, Table 6.

**Table 6.** Thematic analysis of the interview data and selected quotes of the respondents (Theme 5)

| Theme 5 Psychological | Selected quotes |
| --- | --- |
| perspectives<br>(Experience/Outcome) |  |
| **Subthemes** |  |
| 5.1 Fear of COVID-19 | 5.1.1 Contraction<br>*"Yes! I am scared a little bit."* (IMS-3)<br>*"Umm! I am not scared! And if we take precaution, there is no way of transmission of this disease."* (IMS-5)<br>*"I am not that kind of scared but there is a risk, so you must be precautionous."* (IMS-2)<br>5.1.2 Death<br>*It's not that deadly. Its mortality rate is only 2% and 98% of people get recoveries from it.* (IMS-8)<br>5.1.3 Isolation<br>*"It's not enjoyable. It's difficult."* (IMS-7)<br>*"I guess, it is good because no one enters in hostel, campus."* (IMS-9)<br>5.1.4 Effect on career or studies<br>*After the coronavirus pandemic, I have been isolated in my room and even my regular daily activities have been changed drastically like I'm not able to go outside that much and even to my college and hospital everywhere that I used to attend previously in the normal days, that has been hampered in my life.* (IMS-6)<br>*"Oh! I'm making my article. I have 20 articles in my room."* (IMS-9) |
| 5.2 Self-esteem | *"Yea! Due to both."* (IMS-1)<br>*"Ah! No! My self-esteem is not low because I do believe that we have a responsibility for yourselves and society to follow lockdown and carefully follow self-hygiene. If we care about our responsibilities then there are no worries; whether we get infected or not. Because if even we get infected, we know that had tried to follow the guidelines and responsibilities well."* (IMS-11)<br>*"No! Like in the early days, it was kind of enjoyable like we were getting holidays from university. It was relaxing eating but nowadays, as the days are increasing, it's like more loneliness or something like that. We are more alone. We are bored by doing the same thing daily. So, kind of someday sometimes we get frustrated. We are given a single room, so it's not enjoyable."* (IMS-8) |
| 5.3 Loneliness | *"Yes! I am missing them. I miss them so badly that I cried some days like I can't say but the condition is so worse that I can't even go."* (IMS-8)<br>*"Certainly! I am missing my family."* (IMS-2) |
| 5.4 Stress | *"Yes! Lots of changes. It's mentally challenging."* (IMS-1)<br>*"I have never stayed in the room for 2 months as we are right now so I am getting too* |
|  | *much bored.” (IMS-4)*<br>*“Yes! I am sort of yea, I am like, I am scared that what will happen. I am in Pakistan, I am not with my family members.” (IMS-8)* |

*Abbreviations:- COVID-19: Coronavirus disease-2019, IMS 1-11: International medical student 1-11*

##### Contraction

The responses are depicted in Table 6.

##### Death

Though the respondents were scared of being infected, they did not have fear of death, Table 6.

##### Isolation

The mixed views were obtained regarding isolation, Table 6.

##### Effect on career or studies

Though the respondents indicated interruption of regular classes, they were managing time for study in the hostel room and some of them had even started scientific writing, Table 6.

#### Self-esteem

Upon a question related to lowering of self-esteem due to either COVID-19 pandemic or due to lock down or both, mixed reactions were obtained. Some of the respondents stated both as a reason for lowering self-esteem. However, the majority of the respondents reported no lowering at all in their self-esteem as those approaches were adopted for a good reason, Table 6.

#### Loneliness

The majority of the respondents were missing their family members back home, while some were frustrated and facing difficulties, Table 6.

#### Stress

The respondents were found in stress as they were far from family members, staying inside the hostel room for so long, and their minds were filled with the uncertainty of disease progression, Table 6.

## DISCUSSION

The outcome of this study has portrayed the typical picture of international medical students amid the COVID-19 pandemic from several aspects such as; knowledge about COVID-19, perceptions and attitudes towards it, and preparedness and safety against it.

The present study reported that the international medical students had adequate knowledge about the origin, causative agent, nature, and treatment strategy of COVID-19. However, some of them had less information about its treating agents and vulnerability issues. Our findings are in line with the findings of studies conducted in different parts of the world.^14-17^ Whereas the findings from a study conducted in Turkey were somewhat in different line where the knowledge of the medical students was found moderate.^18^ The audio-visual demonstrations, online webinars, and regular capacity development activities help improve the knowledge and skill in such pandemics. COVID-19 pandemic has worryingly endangered the existence of people around the globe. The unavailability of proven therapeutic agents and lockdown and social distancing adopted as preventive measures to contain such pandemic has disturbed the normal pattern of life everywhere.^19,20^

It was identified that the preventive measures were adopted at an adequate level by all the international students. Social distancing, hand washing with soap and water or using sanitizer, wearing masks and gloves, maintaining social distancing, limiting movements, changing and washing clothes frequently, and taking showers were the most frequently adopted preventive measures to avoid infection. Different studies have demonstrated the effectiveness of preventive measures as an appropriate strategy to limit the spread of coronavirus.^6,21^ Similar preventive measures were adopted by medical students in a study conducted by Khasawneh et al in Jordan.^16^ Whereas medical students were least compliant towards the adoption of such preventive approaches in a study conducted by Haque et al.^22^ Preventive measures are, no doubt, helpful due to the unavailability of approved therapeutic agents. But, the regular testing of COVID-19 in mass, effective contact tracing, increasing number of hospital beds, capacity of intensive care units (ICU), ventilators, and effective facility of quarantine and isolations are need of an hour.^23^ Preventive measures coupled with strengthening medical infrastructure throughout the country help ineffective containment of such pandemics.

The finding of the perception of risk of getting infected with COVID-19 indicates the requirement of the counseling sessions and stringent preventive measures. Though the respondents considered themselves safer than other people outside the periphery, most of them felt the risk of transmission from the attendants and other fellow hostelers. Willingness to stay in quarantine or in isolation in case of being exposed to or infected with COVID-19 exhibits a positive attitude. Our findings contrast with the results of a study conducted in Jordan where 4.2% of students were intended to escape isolation in case they were tested COVID-19 positive.^16^ Whereas the outcome of our study is in line with the study conducted by Maheshwari et al in India which demonstrated a positive attitude and appropriate practice in the majority of the participants.

The present study also demonstrates the social nature of students such as; willingness to help society with the COVID-19 pandemic and support friends in case of getting infected. Such an attitude depicts peace of mind. However, the concept of “first me” expressed in some cases might be attributed to the adverse scenario and mental stress caused by such pandemic.

The respondents of the study believe that lockdown is the proper approach for the management of such pandemics. Contrary to this, different studies suggest that the theoretical research used for implementing the lockdown approach was farther than the practical aspect which had neglected the associated psychological effects.^24,25^ Altman M describes lockdown as a misapplied and sub-optimal policy as it has disturbed the world’s economy and emphasizes on the inclusion of more comprehensive governance and improvised mental models as crucial factors for human welfare in such measures.^24^ A study conducted by Zheng et al reported that the lockdown approaches contributed a buffer effect on social anxiety and the mediating effect on psychological distancing.^25^

The eagerness of international students to get vaccinated, as soon as it is available, implies their willingness to be stress-free of the pandemic and trust towards the immediate development of scientific research. However, at the same time, some of them were in a wait-and-see mood for the same. A similar type of result was reported by various studies where the majority of the people were willing but some were either unwilling or uncertain to get vaccinated.^26-29^ The reluctant respondents in the studies were mostly from ethnic minorities and had lower levels of education and income.^26,29^ Different reports suggest that the mistrust towards vaccines and fear of future side effects are responsible for people’s reluctance towards getting a vaccination. ^26,30,31^ Doubt in the current treatment strategy being adopted as a therapeutic option is creating fear among students. Such poor compliance urges for an immediate policy response tailored to public health awareness programs.^26^

The present study identified mixed responses from international students about the university and hostel management towards their welfare amid the COVID-19 pandemic. The majority of the respondents reported a satisfactory response, however, some of them were found a little bit dissatisfied with the authority. The respondents said that their normal routine has been disturbed due to the university shut down, restriction in movement, and confinement within the hostel periphery. In addition to this lack of physical activity, mess closure, the uncertainty of the situation make them miss their family more which may lead to psychosocial stress. Opening of mess for international students, proper hygiene and sanitation measures, virtual teaching-learning process, and online awareness training and webinar regarding the containment aspect of disease to protect themselves and to the society is required. School closure and loss in their learning are some of the prime concerns raised by the respondents. Lockdown has affected the global education system. The education system in LMICs was already facing many challenges, COVID-19 adds more stress to it. This establishes the urgent need for multimodal approaches for achieving the course content objectives enabling learners in analytical reasoning to deal with the complexity of online education.^32^ Timely and significant teaching-learning techniques must be implemented for medical students as it is mandatory to have a significant understanding and education that enhances performances in such pandemics for the safety of the country.^18^

The study reports the compliance of respondents with the COVID-19 containment strategies of the government. The safety and security concerns, opening groceries for international students for a limited time, medical facilities, as well as regular and authentic sharing of COVID-19 information through newspapers, are praiseworthy steps taken government. The majority of respondents’ intact self-esteem even at a bit frustrating scenario like this implies their intact mental status as well as good governance.

Multiple stressors may contribute to psychological stress and anxiety such as the perceived risk of infection, reduced social communications, and augmented worries on academic performance.^33^ The study also depicts the same. Most of the respondents had fear of getting contracted and impact in study and career but nobody had any fear of death as the mortality rate of COVID-19 is very low as compared to many other common diseases.^34^ A study conducted by Saddik et al reported higher anxiety levels in medical students during clinical rotation that decreased with online learning.^15^ Crisis like the COVID-19 pandemic gives substantial psychosocial burdens on humankind. Therefore, developing interventions and preventive strategies are crucial to address the psychological impact on international students due to such a pandemic.^33^ The focus must be given towards solidarity and social justice to deal with social stigma as well, which may occur at any point amid such pandemic and, if go unchecked, may lead to the mental disorder.^35^ A proper health communication is an urgent requirement as per WHO’s the current report for the quality of life both from the health and social point of view.^36^ Cohesiveness and solidarity are effective coping mechanisms for stress and anxiety.^33^

Therefore, there is a need for an immediate action plan, supported by behavioral and social sciences, for alleviating the possibly overwhelming impacts of such pandemics to help align human behavior.^36^

This study depicts the compliance of international students with the guidelines for effective management of the COVID-19 pandemic. The finding of the study aligns with the study by Saddik et al where medical students were found to be utilizing reliable sources of information for COVID-19 management.^15^ Such adherence is critical for the effective management of such a global health threat. Lifestyle adopted for well-being amid such a pandemic by international students were timely and productive as well. Many of them were engaged in scientific publications which was a quality fruitful way to live for medical students.

### Strengths and limitations

This study is novel as it is the first of its kind study. It has delivered a convenient and comprehensive understanding of the knowledge, perception, and preparedness of the international medical students residing in hostels of different Universities of Pakistan towards the COVID-19 pandemic. A significant strength of this study was the inclusion of international students of six countries who are pursuing medical courses in seven Universities in different places of Pakistan. This enabled an in-depth exploration of the range of knowledge, perceptions, and preparedness as well of such students towards the COVID-19 pandemic. This study informs about the status of international medical students in Pakistan that can help their management with a quality stay in such a global pandemic. In this context, this study can have an impact in filling the prevailing gap in the qualitative, academic, and administrative issues between the international medical students and the respective authority and the literature as well. All the participants were guaranteed the secrecy and confidentiality of their profile, and the interview was also taken by international students in Pakistan. Therefore, the participants liberally and decently communicated their feelings. Besides, the data analysis based on a thematic analysis method simplified the generation of typical findings.

Moreover, there are some probable limitations linked to this qualitative study. This study was carried out only in the universities located in the Punjab province, Pakistan. Therefore, the findings of this study may not be generalized to the whole country, but the findings in other provinces may not vary with our findings.

## CONCLUSION

The study furnishes important information on the perception and preparedness of international medical students regarding the COVID-19 pandemic. Respondents believed that maintaining personal hygiene using masks, gloves, and sanitizers and frequent hand-washing; maintaining social distancing; and complying with lockdown measures aid in COVID-19 management. The majority of them had a clear understanding of the COVID-19 pandemic. The support from the universities is the primary requirement for international students to cope with such pandemic as there is no approved therapy and preventive measures cannot always be adequate to sufficiently thwart COVID-19. Furthermore, the chance of psychological stress is high that could substantially harm their quality of life. The findings from the present study demand timely and evidence-based teaching-learning techniques for medical students as it is mandatory to have a significant understanding that enhances performances in such pandemics for global safety. Such a technique could ensure their mental health and make them self-motivated. In such a situation, the students prefer to use their time productively and this could have a long-term effect on their careers and academic progress. The respondents’ perspectives from the present study help government to prepare relevant public health strategies based on population-focused approaches for effective management of such a disaster.

## Data Availability

There are no data in this work.

## Author contributions

Conceptualisation: SK, MU. Methodology: SK, MU, RPG, YB, and JS. Formal analysis: SK, MU, MS, HR, and MA. Writing-original draft: SK, SG, and MU. Writing-review and editing: MS, HR, SG, and HAS. Approval of final manuscript: all authors.

## Special acknowledgment

We are highly grateful to Mr. Tank Prasad Yadav and Mr. Pallav Aryal for their support in the finalization of this manuscript.

## Ethical approval

The ethics approval was received from the Institutional Ethics Review Board of the University of the Punjab, Lahore, Pakistan (Reference number: 183/DFEMS). Informed verbal consent to participate in the study was obtained from participants.

